# Association of Urine Findings with Metabolic Syndrome Traits in a Population of Patients with Nephrolithiasis

**DOI:** 10.1101/2021.03.26.21254406

**Authors:** Virginia L. Hood, Kevan M. Sternberg, Desiree de Waal, John R. Asplin, Carley Mulligan, Peter W. Callas

## Abstract

**Background and objectives:** The odds of nephrolithiasis increase with more metabolic syndrome (met-s) traits. We evaluated associations of metabolic and dietary factors from urine studies and stone composition with met-s traits in a large cohort of stone-forming patients.

**Design, setting, participants & measurements:** Patients >18 years, who were evaluated for stones with 24 h urine collections, July 2009-December 2018, had records reviewed retrospectively. Patient factors, laboratory values and diagnoses were identified within 6 months of urine collection and stone composition within 1 year. Four groups with 0, 1, 2, ≥ 3 met-s traits (hypertension, obesity, dyslipidemia, diabetes) were evaluated. Trends across groups were tested using linear contrasts in analysis of variance and analysis of covariance.

**Results:** 1473 patients met inclusion criteria (835 with stone composition). Met-s groups were 0=684, 1=425, 2=211, 3 and 4 =153. There were no differences among groups for urine volume, calcium or ammonium (NH4) excretion. There was a significant trend (p<0.001) for more met-s traits being associated with decreasing urine pH, increasing age, calculated dietary protein, urine uric acid, oxalate, citrate, titratable acid (TAP), net acid excretion (eNAE) and uric acid supersaturation. The ratio of ammonium to net acid excretion did not differ among the groups.

After adjustment for protein intake, the fall in urine pH remained strong, while the upward trend in TAP excretion was attenuated and NH4 decreased. Calcium oxalate stones were most common, but there was a trend for more uric acid (p<0.001) and fewer calcium phosphate (p=0.09) and calcium oxalate stones (p=0.01) with more met-s traits.

**Conclusions:** Stone forming patients with met-s have a defined pattern of metabolic and dietary risk factors that contribute to an increased risk of stone formation including higher acid excretion, largely the result of higher protein intake, and lower urine pH.

## INTRODUCTION

Metabolic syndrome (met-s) with its components of insulin resistance, obesity, dyslipidemia and hypertension, is a major health challenge increasing cardiovascular risk (1, 2) and the risk for kidney stones (2-4). The odds of nephrolithiasis increases with the number of met-s traits (5, 6).

The standard American dietary pattern, characterized by excess sugar, refined carbohydrates, red and processed meats, pre-packaged foods, high fat fried foods and high sugar beverages, is prevalent amongst individuals with met-s (7). This dietary pattern is associated with increased acid production. The kidneys respond to this acidogenic diet by increasing net acid excretion (NAE), excretion of sulfate, phosphate, urate, chloride, calcium, organic anions forming titratable acid and ammonium plus reduced excretion of bicarbonate and citrate (8). The formation of kidney stones is multifactorial and associated with varying concentrations of these urine components that serve as constituents or inhibitors of stone formation along with urine pH. A low urine pH enhances the formation of uric acid, and to a lesser extent calcium oxalate stones, both of which are common in those with met-s (9-11). Prior studies in non-stone forming persons have shown an inverse relationship between the number of met-s traits and urine pH as well as acid excretion patterns with a lower proportion of ammonium excretion to net acid excretion being associated with more met-s traits (9). The low urine pH in those with met-s has been postulated to result from insulin resistance with resulting impaired ammonia production (10, 11). Uric acid stone formers with similar net acid excretion to non-stone formers have also been reported to have a lower proportion of ammonium and higher proportion of titratable acid than BMI matched controls (11). We evaluated a large cohort of stone-forming patients to identify patient, urine factors and stone composition associated with varying numbers of met-s traits and in particular, those reflecting acid excretion and diet.

## METHODS

### Study design and participants

(Figure 1) A retrospective review was performed of 24-hour urine studies from patients seen for kidney stone consultation at the University of Vermont Medical Center (UVMMC) from July 2009 to December 2018. Results for patients under the age of 18 and those with improper collections were excluded. Patients with suspected renal tubule acidosis based on urine pH and serum bicarbonate were also excluded. Patient variables, laboratory values, associated diagnoses, and medications were recorded for the time closest to and within 6 months of the of the first available urine collection after 2009 and for stone composition within 1 year of urine collection. Patients were divided into 4 groups based on the number of met-s traits (hypertension, obesity, dyslipidemia, diabetes) 0, 1, 2, 3 & 4. Each trait was defined by established criteria (1).

**FIGURE 1.**
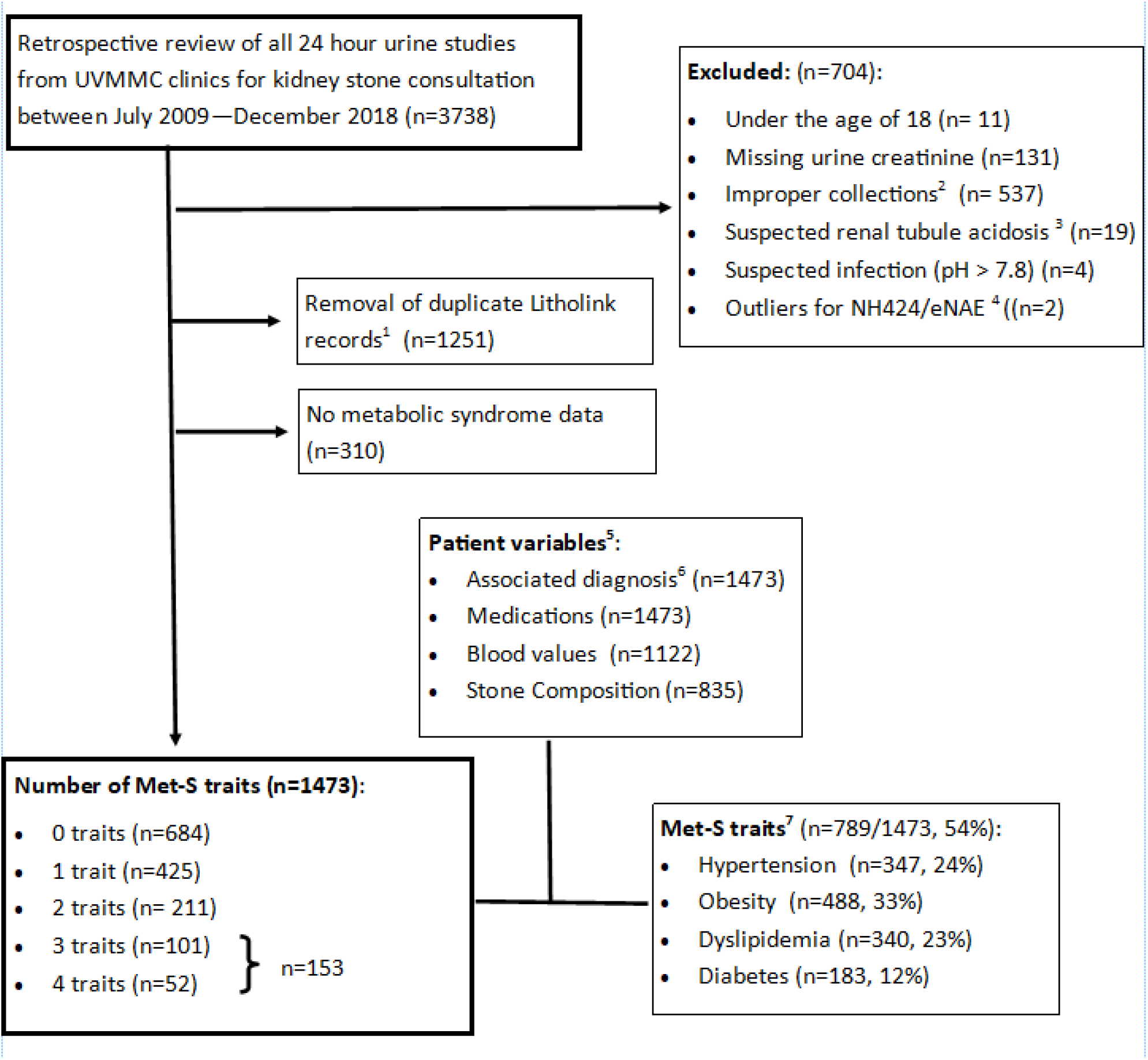
STUDY FLOW DIAGRAM. 1. If a patient had more than one urine test, the earliest one obtained was used to minimize any treatment effects 2. Improper collections defined as outside expected range of urine creatinine based on weight (8.7-20.3 mg/kg for females; 11.9-24.4 mg/kg for males) using Litholink range as reference. Urine creatinine values prior to 4/29/18 were adjusted to account for a change to an IDMS traceable urine creatinine assay 3. Renal tubule acidosis defined as blood tCO2 <22 and urine pH > 6.5 4. Outliers defined as NH424/eNAE absolute values of 194 and 74 with next highest=16 5. Blood values within 6 months of urine collection; Stone composition values within 1 year of urine collection 6. Diagnosis based on ICD 9 and 10 codes listed on “problem list” in medical records 7. Met-S traits defined (1) as needing at least one quality listed: • Hypertension defined as: Diagnosis on problem list, 2 systolic BP readings > 140 mmHg within 6 months of one another, on at least 2 medications for hypertension • Obesity defined as: Diagnosis on problem list, BMI > 30 kg/m2 • Dyslipidemia defined as: Diagnosis on problem list; Triglyceride level > 150 mg/dl; HDL < 35; On a “statin” • Diabetes defined as: Diagnosis of diabetes or hyperglycemia on problem list; Random blood sugar > 180 mg/dl; on oral hypoglycemic agent or insulin

### Data Collection and measurement

Blood levels were measured in UVMMC laboratory by standard autoanalyser methods. 24 hour urine values were measured through Litholink® (Chicago, IL). These included pH, supersaturation of calcium oxalate(SSCaOx), supersaturation of calcium phosphate(SSCaP), supersaturation of uric acid(SSUA) and 24 hour excretion rates for volume (liters/day),calcium(Ca) mg/day, oxalate(Ox) mg/day, citrate(Cit) mg/d, uric acid(UA) g/d sodium(Na) mmol/day, potassium(K) mmol/day, magnesium(Mg)mg/day, phosphorus(P) g/day, chloride(Cl) mmol/day, ammonium(NH4) mmol/d, sulfate(Sul) mEq/d, urine urea nitrogen(UUN) g/day, creatinine mg/day, weight (Kg).

Stone composition was analyzed by Mayo clinic laboratories in those who had passed stones or had them removed within a year of the urine collection. Many stones had several components. For the analysis, stones were categorized as greater than 50% of either calcium oxalate, calcium phosphate, uric acid or other.

### Calculations

(Table 1) Net acid excretion (eNAE) was calculated as sum of urine ammonium and calculated titratable acid from phosphate (TAP) minus calculated bicarbonate, in mEq/day. Urine organic acids were not measured and thus titratable acid from organic acids could not be assessed. TAP accounts for more than 75% of titratable acid (12). Protein intake in grams/day was estimated from UUN and non-urea nitrogen (13). GI alkali intake (mEq/day) was estimated from the difference between excretion of urine cations (Na, K, Ca, Mg) and anions (Cl, P) (14).

**Table 1:**
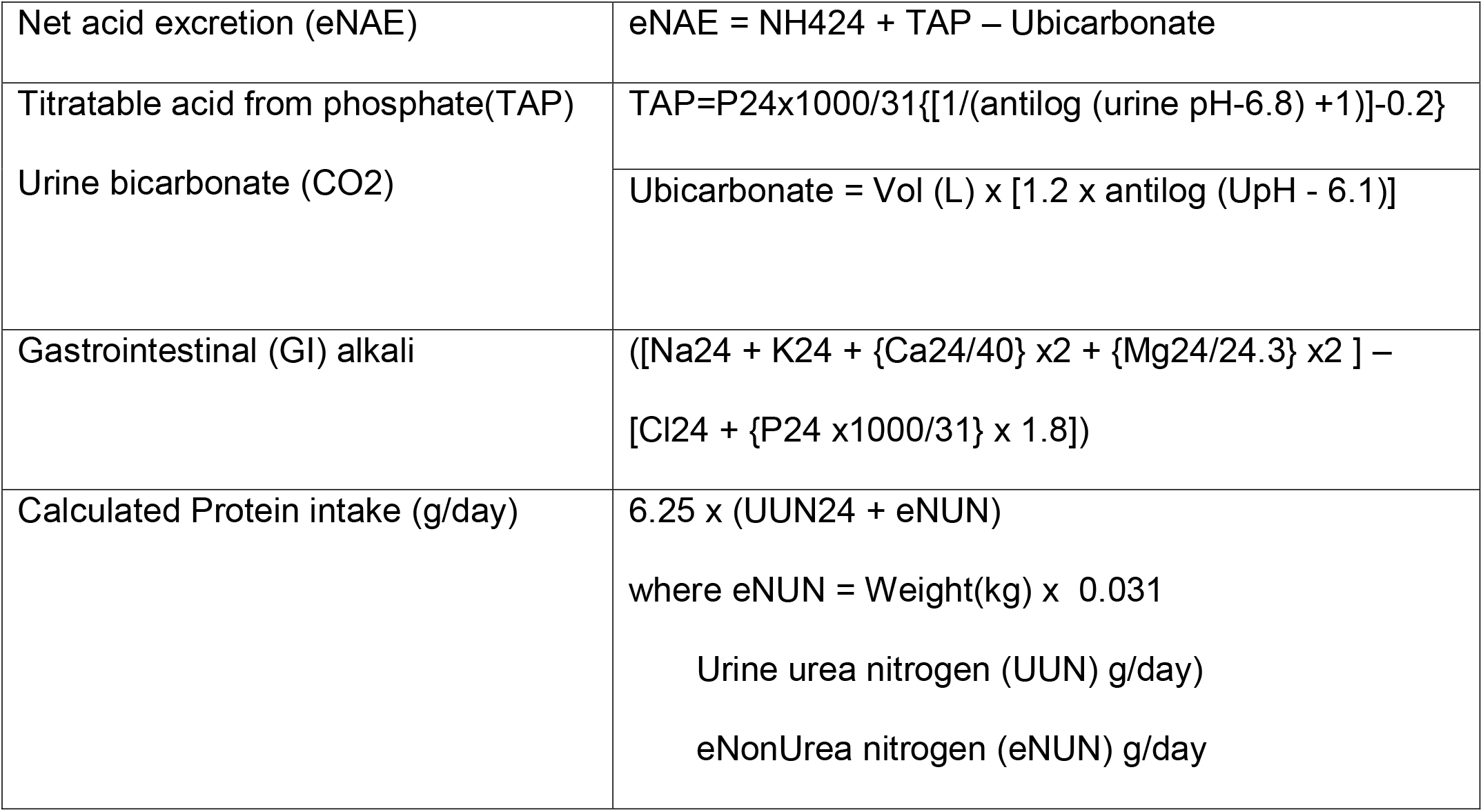
Equations and Calculations.

The information analyzed was collected as part of clinical care, housed in the Stone Registry at UVMMC and extracted in de-identified formats for analysis. This registry is updated yearly and is approved by UVM IRB for use in studies that fit acceptable criteria.

### Statistical Analysis

Analysis was conducted using SAS 9.4 (Cary, NC:SAS Institute Inc.)

For unadjusted comparisons, trends across the four met-s groups were tested using linear contrasts in analysis of variance for continuous variables and Cochran-Armitage trend tests for categorical variables. Adjusted comparisons used linear contrasts in analysis of covariance.

## RESULTS

Results are shown in Tables 2-4. 1473 unique patients met criteria for inclusion. The number of patients in each met-s category was: 0=684, 1=425, 2=211, 3 or more =153 (101 with 3, 52 with 4). Those with more met-s traits were heavier and had higher estimated protein intake (Tables 2 and 3).

**Table 2:**
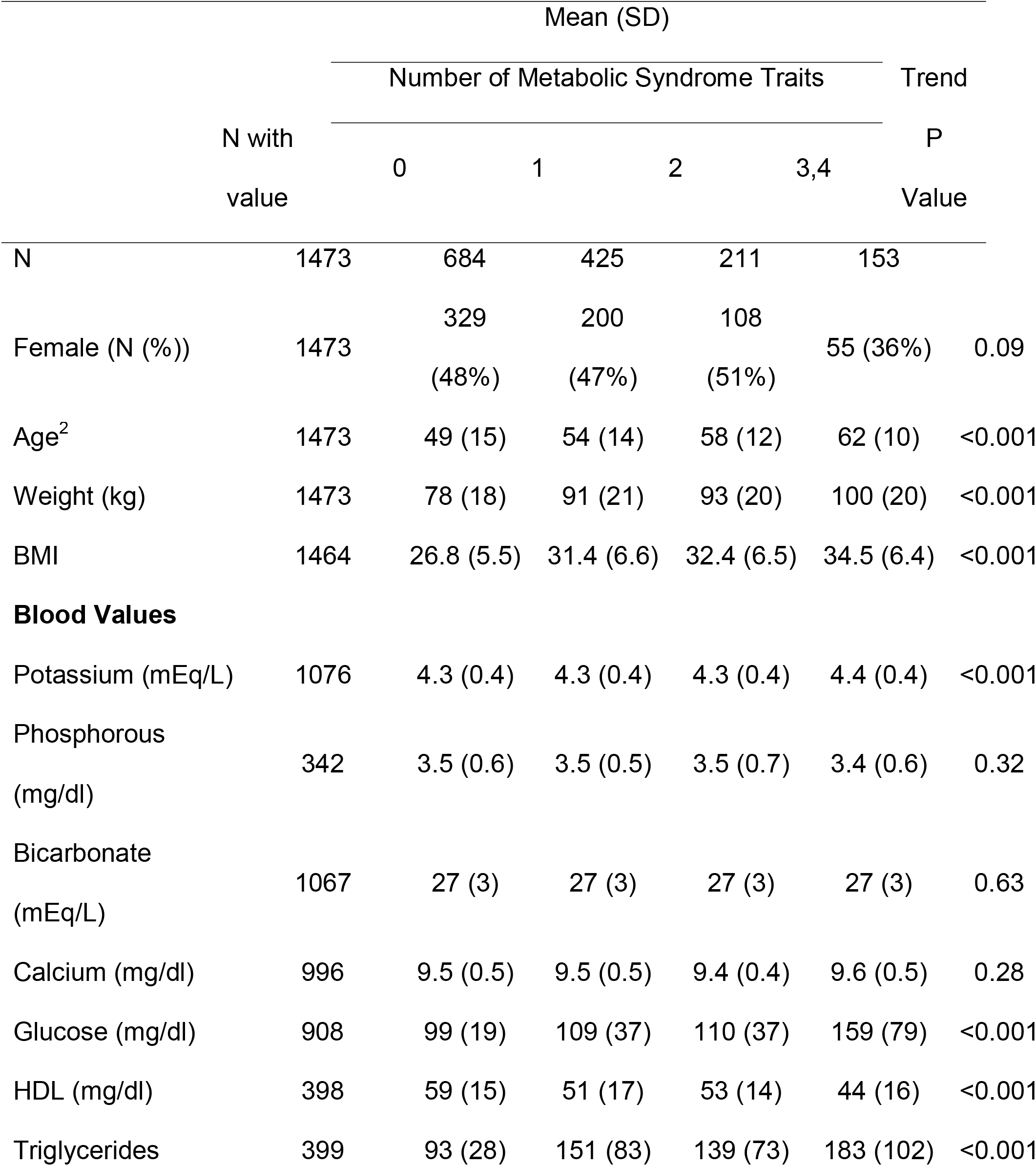
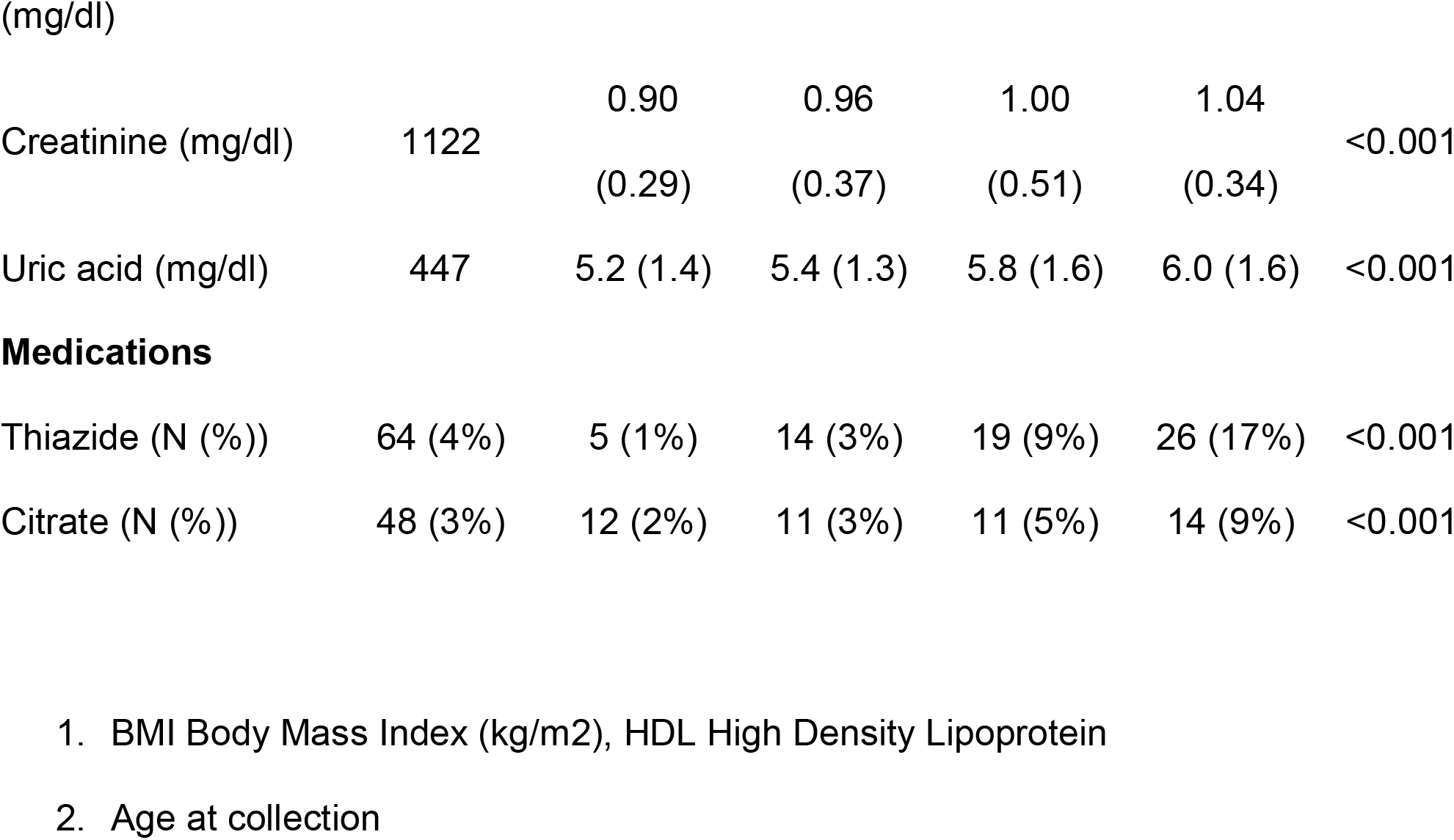
Demographics, Medications and blood values^1^.

**Table 3.**
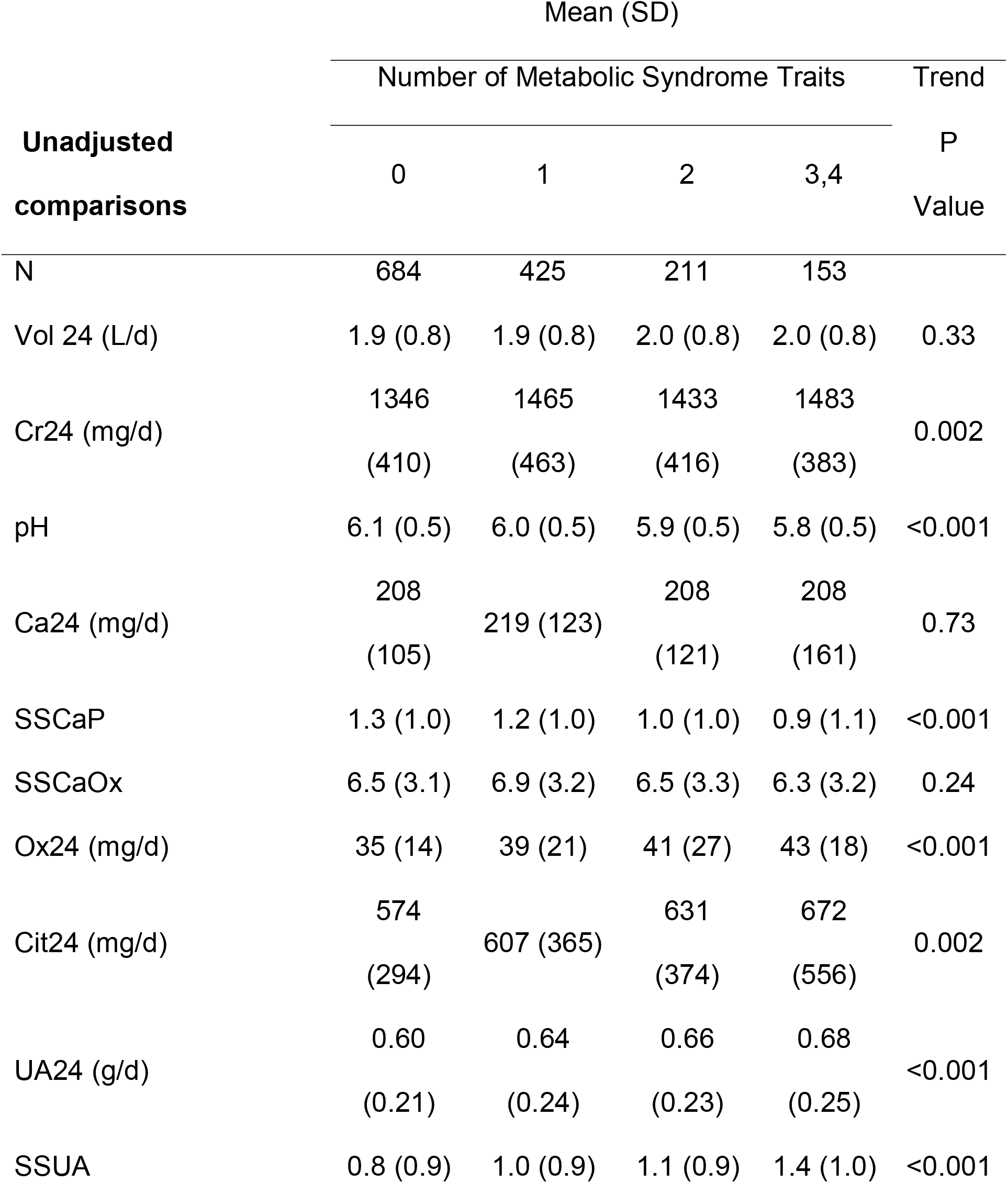
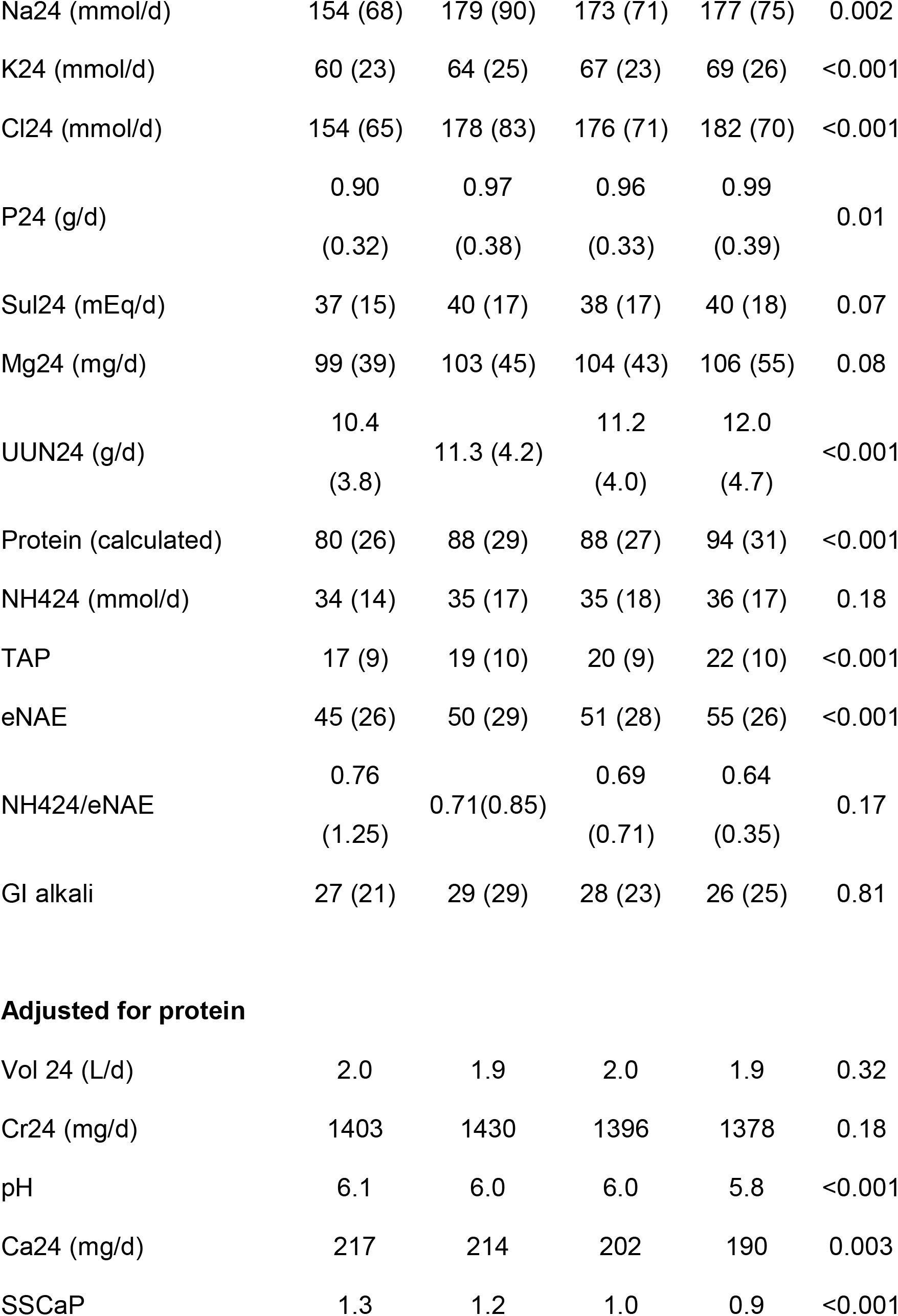
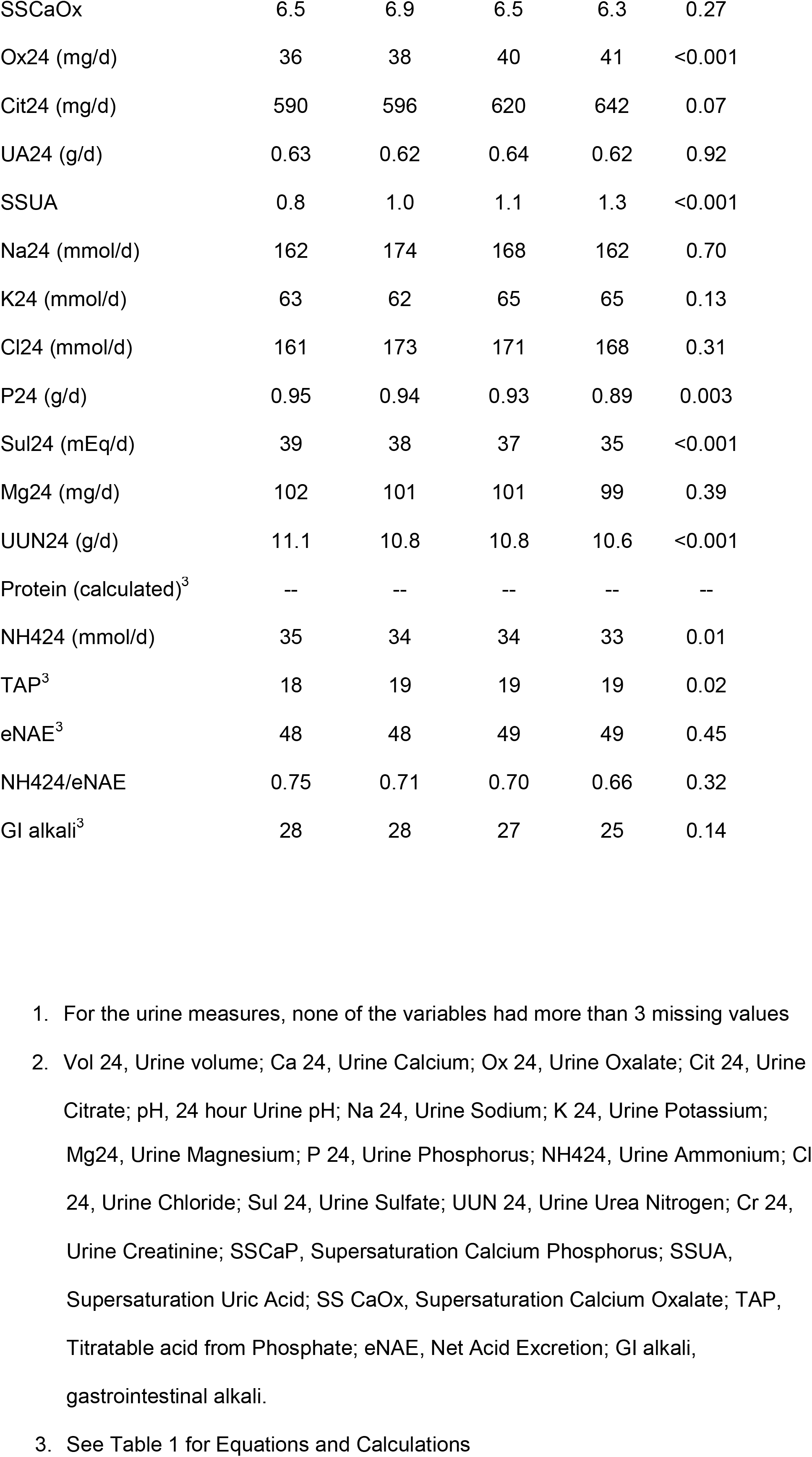
Comparison of Urine Variables and Metabolic Syndrome Traits ^1,2^.

| Unadjusted<br>comparisons | Mean (SD) |  |  |  | Trend<br>P<br>Value |
| --- | --- | --- | --- | --- | --- |
|  | Number of Metabolic Syndrome Traits |  |  |  |  |
|  | 0 | 1 | 2 | 3,4 |  |
| N | 684 | 425 | 211 | 153 |  |
| Vol 24 (L/d) | 1.9 (0.8) | 1.9 (0.8) | 2.0 (0.8) | 2.0 (0.8) | 0.33 |
| Cr24 (mg/d) | 1346<br>(410) | 1465<br>(463) | 1433<br>(416) | 1483<br>(383) | 0.002 |
| pH | 6.1 (0.5) | 6.0 (0.5) | 5.9 (0.5) | 5.8 (0.5) | <0.001 |
| Ca24 (mg/d) | 208<br>(105) | 219 (123) | 208<br>(121) | 208<br>(161) | 0.73 |
| SSCaP | 1.3 (1.0) | 1.2 (1.0) | 1.0 (1.0) | 0.9 (1.1) | <0.001 |
| SSCaOx | 6.5 (3.1) | 6.9 (3.2) | 6.5 (3.3) | 6.3 (3.2) | 0.24 |
| Ox24 (mg/d) | 35 (14) | 39 (21) | 41 (27) | 43 (18) | <0.001 |
| Cit24 (mg/d) | 574<br>(294) | 607 (365) | 631<br>(374) | 672<br>(556) | 0.002 |
| UA24 (g/d) | 0.60<br>(0.21) | 0.64<br>(0.24) | 0.66<br>(0.23) | 0.68<br>(0.25) | <0.001 |
| SSUA | 0.8 (0.9) | 1.0 (0.9) | 1.1 (0.9) | 1.4 (1.0) | <0.001 |
| Na24 (mmol/d) | 154 (68) | 179 (90) | 173 (71) | 177 (75) | 0.002 |
| K24 (mmol/d) | 60 (23) | 64 (25) | 67 (23) | 69 (26) | <0.001 |
| Cl24 (mmol/d) | 154 (65) | 178 (83) | 176 (71) | 182 (70) | <0.001 |
| P24 (g/d) | 0.90<br>(0.32) | 0.97<br>(0.38) | 0.96<br>(0.33) | 0.99<br>(0.39) | 0.01 |
| Sul24 (mEq/d) | 37 (15) | 40 (17) | 38 (17) | 40 (18) | 0.07 |
| Mg24 (mg/d) | 99 (39) | 103 (45) | 104 (43) | 106 (55) | 0.08 |
| UUN24 (g/d) | 10.4<br>(3.8) | 11.3 (4.2) | 11.2<br>(4.0) | 12.0<br>(4.7) | <0.001 |
| Protein (calculated) | 80 (26) | 88 (29) | 88 (27) | 94 (31) | <0.001 |
| NH424 (mmol/d) | 34 (14) | 35 (17) | 35 (18) | 36 (17) | 0.18 |
| TAP | 17 (9) | 19 (10) | 20 (9) | 22 (10) | <0.001 |
| eNAE | 45 (26) | 50 (29) | 51 (28) | 55 (26) | <0.001 |
| NH424/eNAE | 0.76<br>(1.25) | 0.71(0.85) | 0.69<br>(0.71) | 0.64<br>(0.35) | 0.17 |
| GI alkali | 27 (21) | 29 (29) | 28 (23) | 26 (25) | 0.81 |
| **Adjusted for protein** |  |  |  |  |  |
| Vol 24 (L/d) | 2.0 | 1.9 | 2.0 | 1.9 | 0.32 |
| Cr24 (mg/d) | 1403 | 1430 | 1396 | 1378 | 0.18 |
| pH | 6.1 | 6.0 | 6.0 | 5.8 | <0.001 |
| Ca24 (mg/d) | 217 | 214 | 202 | 190 | 0.003 |
| SSCaP | 1.3 | 1.2 | 1.0 | 0.9 | <0.001 |
| SSCaOx | 6.5 | 6.9 | 6.5 | 6.3 | 0.27 |
| Ox24 (mg/d) | 36 | 38 | 40 | 41 | <0.001 |
| Cit24 (mg/d) | 590 | 596 | 620 | 642 | 0.07 |
| UA24 (g/d) | 0.63 | 0.62 | 0.64 | 0.62 | 0.92 |
| SSUA | 0.8 | 1.0 | 1.1 | 1.3 | <0.001 |
| Na24 (mmol/d) | 162 | 174 | 168 | 162 | 0.70 |
| K24 (mmol/d) | 63 | 62 | 65 | 65 | 0.13 |
| Cl24 (mmol/d) | 161 | 173 | 171 | 168 | 0.31 |
| P24 (g/d) | 0.95 | 0.94 | 0.93 | 0.89 | 0.003 |
| Sul24 (mEq/d) | 39 | 38 | 37 | 35 | <0.001 |
| Mg24 (mg/d) | 102 | 101 | 101 | 99 | 0.39 |
| UUN24 (g/d) | 11.1 | 10.8 | 10.8 | 10.6 | <0.001 |
| Protein (calculated)<sup>3</sup> | -- | -- | -- | -- | -- |
| NH424 (mmol/d) | 35 | 34 | 34 | 33 | 0.01 |
| TAP<sup>3</sup> | 18 | 19 | 19 | 19 | 0.02 |
| eNAE<sup>3</sup> | 48 | 48 | 49 | 49 | 0.45 |
| NH424/eNAE | 0.75 | 0.71 | 0.70 | 0.66 | 0.32 |
| GI alkali<sup>3</sup> | 28 | 28 | 27 | 25 | 0.14 |

1. For the urine measures, none of the variables had more than 3 missing values 2. Vol 24, Urine volume; Ca 24, Urine Calcium; Ox 24, Urine Oxalate; Cit 24, Urine Citrate; pH, 24 hour Urine pH; Na 24, Urine Sodium; K 24, Urine Potassium;
Mg<sub>24</sub>, Urine Magnesium; P<sub>24</sub>, Urine Phosphorus; NH<sub>4</sub><sub>24</sub>, Urine Ammonium; Cl<sub>24</sub>, Urine Chloride; Sul<sub>24</sub>, Urine Sulfate; UUN<sub>24</sub>, Urine Urea Nitrogen; Cr<sub>24</sub>, Urine Creatinine; SSCaP, Supersaturation Calcium Phosphorus; SSUA, Supersaturation Uric Acid; SS CaOx, Supersaturation Calcium Oxalate; TAP, Titratable acid from Phosphate; eNAE, Net Acid Excretion; GI alkali, gastrointestinal alkali.
3. See Table 1 for Equations and Calculations

### Blood values : (Table 2)

1122 had at least one value for blood chemistries within 6 months of the Litholink. There was a significant trend for increasing glucose, TG and decreasing HDL with increasing numbers of met-s traits as would be expected, given the defining criteria. There were upward, though quantitatively small, trends for serum potassium and creatinine. Uric acid also showed an upward trend as previously described in those with met-s (10, 11). There were no differences among the groups for calcium, phosphorus or total CO2.

### Medications: (Table 2)

Of the 1473 patients, 64 (4%) had a thiazide (either alone or combination with ACEi or ARB) noted with a trend to a greater proportion in those with more met-s traits p<0.001); 48 (3%) had a citrate (potassium, sodium or combination) noted, also with a trend to a greater proportion in those with more met-s traits (p< 0.001).

### Urine values: (Table 3, Table 1S)

There were no significant trends for unadjusted values for the groups for 24 hour urine volume, calcium, sulfate, magnesium, SSCaOx, NH4 or NH4/eNAE. There was a significant trend for the increasing number of met-s traits being associated with decreasing urine pH and decreasing SSCaP and increasing urine UA, SSUA, oxalate, citrate, Na, K, P, UUN, TAP and eNAE.

When adjusted for calculated protein intake or age and protein intake, the trend for decreasing urine pH remained strong as did the upward trend for oxalate. However the upward trend for TAP was attenuated, and there was no longer a significant trend for eNAE, Na, K, or citrate. There was a significant downward trend for UUN, phosphorus and sulfate. When adjusted for age NH4 showed an upward trend (Table 1S, p<0.001) but when adjusted for protein intake there was a downward trend (Table 3, p<0.01) and in combination, no trend (Table 1S, p=0.93).

The ratio of NH4/eNAE did not significantly differ among the groups.

The calculated value for GI alkali (Table 3,Table 1S) showed a downward trend after adjusting for age and protein (p< 0.01) but not for either age or protein alone.

### Stone composition

(Table 4) 835 (57%) had information about stone composition within a year of the Litholink values – 593 (71%) had > 50% calcium oxalate, 126 (15%) > 50% calcium phosphate, 83 (10%) had > 50% uric acid and 33 (4%) mixed/other stones (see Table 4 for composition). Although calcium oxalate stones were the most common in all groups, there was a trend for an increasing proportion of uric acid stones (p< 0.001) and decreasing proportion of calcium phosphate stones (p=0.09) and calcium oxalate stones (p=0.01) with an increasing number of met-s traits.

**Table 4:** Predominant Stone Composition.

| Stone comp | 0 criteria | 1 criteria | 2 criteria | 3 ,4 criteria | P value |
| --- | --- | --- | --- | --- | --- |
| N = 802* | 376 | 222 | 112 | 92 |  |
| Uric acid | 24 (6%) | 18 (8%) | 11 (10%) | 30 (33%) | <0.001 |
| CaP | 67 (18%) | 37 (17%) | 15 (13%) | 7 (8%) | 0.09 |
| CaOx | 285 (76%) | 167 (75%) | 86 (77%) | 55 (60%) | 0.01 |
| Stones with any uric acid composition |  |  |  |  |  |
| N=835 | 398 | 229 | 116 | 92 |  |
| Any uric acid | 28 (7%) | 20 (9%) | 12 (10%) | 30 (33%) | <0.001 |

\* 835 patients had stone data, 2 were removed with predominately infection stones, 2 with predominately cystine stone, 1 with predominately other (CaCarb stone). 28 of these patients did not have >50% of any one stone constituent and were removed from this data.

## DISCUSSION

Our study shows that in a large group of stone forming persons with varying numbers of met-s traits, urine pH and acid excretion patterns were similar to those described by others in non-stone forming persons with met–s (9). However, the changes associated with met-s in our study were largely the consequence of higher dietary protein intake. Those with more met-s traits had higher weights and ate more protein which, except for urine pH, accounted for most of the variability in acid excretion findings among those with varying numbers of met-s traits. Other urinary findings of metabolic and dietary factors contributing to excess stone formation in those with met-s were also influenced by diet as discussed below.

### Acid excretion

The urine metabolic profiles showed similar acid excretion findings to a previously published study of a smaller group of non-stone forming persons with and without met-s traits (9). The main findings in both this and our study were that with an increasing number of met-s traits, there was decreasing urine pH and greater net acid excretion (eNAE) that was contributed to by both a greater ammonium (age adjusted) and titratable acid excretion. However, in contrast to the referenced study, we did not show a significant decrease in the ratio of NH4/eNAE and thus cannot ascribe the change in acid excretion to a shift away from ammonium excretion (9).

The main influences on the acid excretion variables in the met-s groups was protein intake and age. When adjusted for protein or combination of these factors, the increases in ammonium and net acid excretion were attenuated notably.

Urine pH is the only variable that was not influenced by either age or protein intake or any other urine factors analyzed, and is independently associated with met-s traits in this analysis as has been noted by others. (9, 10)

Our findings suggest that those with more met-s traits have higher weights and consume more protein and that the changes in renal acid excretion are heavily influenced by dietary protein intake. In the previous study of non-stone formers with met-s, where NAE depended relatively less on ammonium excretion and more on TA excretion than in those without met-s, diet was not controlled and there was no mention of UUN or estimated protein intake which may have been different among the groups.

Other dietary factors that affect acid excretion patterns are reflected in the amount of estimated GI alkali, a surrogate for alkali intake, and sulfate a surrogate for acidogenic components of animal protein. In the current study, when adjusted for protein intake GI alkali intake trended downward. There was also a downward trend for urine sulfate when adjusted for protein intake with lower values in those with more met-s traits. All these findings reflect dietary influences as those with more met-s traits consume more protein, particularly animal protein with associated less alkali.

### Urine pH

In all prior studies of non-stone formers, urine pH trended down as the number of met-s traits increased (9, 10). This trend was not changed when adjusted for protein in our study. Low urine pH can result from increased acid excretion or reduced urinary buffering or both. Reduced NH4/NAE was noted in a diet controlled study of uric acid stone formers who tend to have a lower urine pH, indicating the low urine pH in these subjects could be due to lower buffering from less NH4 but no comment was made about met-s characteristics (11). We did not find the same pattern in our stone forming patients with a variety of stone compositions and there were too few with predominantly uric acid stones to evaluate them separately. Urine buffers with organic anions were not assessed but as mentioned would likely be quantitatively too small to explain any variability among the groups with respect to the variability in urinary pH.

Increased acid excretion results from increased acid generation that in steady state is largely due to dietary protein intake. The findings of a downward trend for sulfate excretion, a surrogate for animal protein, loss of the upward trend for P excretion and attenuation of the trend for TAP when adjusted for protein could be largely a consequence of differences in dietary animal protein intake, suggesting that those with most met-s traits eat more animal protein. Interestingly, sodium excretion which also trended upward in those with more met-s traits, was lost when adjusted for protein. Acid excretion values were not affected when adjusted for GI alkali. (Table 1S)

Both lower and higher citrate excretion have been previously described in those with met-s (15, 16). We showed an upward trend for citrate excretion in those with more met-s traits that was lost when adjusted for protein intake. Low urine pH and higher acid excretion in those with more met-s traits would reduce not increase citrate excretion.

### Effects of medications

There was an upward trend for both thiazide and citrate intake in those with more met-s traits however it is unlikely that these significantly influenced the urine findings. For urine citrate, taking thiazides would not enhance citrate excretion as thiazides cause hypocitraturia. The percent of patients taking citrate medication was small and although citrate intake could increase its excretion, the quantitative impact is hard to calculate and the effect was lost when adjusted for protein intake making diet rather than medications the likely explanation for the trend to higher citrate excretion in those with more met-s traits. The findings for urine acid excretion values (pH, NH4, NAE, citrate) were not changed by excluding the 53 persons with mention of citrate supplements in their records.

The higher NH4, TAP, eNAE and lower urine pH should not result from the intake of either of these classes or drugs as they both lead to a systemic alkalosis so would not explain the increase of in acid excretion with increased met-s traits.

### Effects of diet

In steady state, urine excretion of Na, K, Ca, P, Mg, UUN (protein surrogate), and volume reflects intake and absorption.

There were trends to higher sodium and phosphorus that were lost and sulfate excretion that decreased when corrected for protein intake. Calcium excretion while not differing in the unadjusted or age adjusted analyses, did decrease when adjusted for protein. Urine potassium excretion was higher in those with more met-s traits in the unadjusted analysis but this trend was lost when adjusted for protein.

Dietary factors in the met-s groups can influence many of the metabolic risk factors found in the urine of stone formers with met-s. Protein intake seems to be the dominant force for the changes noted in urine Ca, P, UA, K, sulfate and citrate but also for Na. As Na and these other factors are highly correlated with protein (Table 2S), the findings after adjustment for protein suggest that those with more met-s traits eat protein associated with more processed food with more Na and P. Only oxalate was not modified in any of the analyses.

**Stone formation** is usually not caused by a single chemical anomaly but rather a combination of factors having an impact on urine super-saturation or loss of inhibitors. Standard American dietary patterns are low in plant-based foods and high in animal protein and additives such as P and Na. Those with more met-s traits appeared to consume more animal protein which tends to increase risk of stone formation due to an interplay of factors including increasing net endogenous acid production.

We only had stone composition information for 57% of our patients and do not know whether these findings represent stones for the entire group. The majority of stones were CaOx with fewer being CaP or UA which is similar to the general population of stone formers. However the pattern of stone composition was similar to that suggested in other studies of met-s with a significant trend to more uric acid stones in those with more met-s traits (10, 17). This is most likely due to the trend to lower urine pH which predisposes to the formation of insoluble uric acid in place of more soluble sodium urate as pH decreases. In addition, both blood and urine uric acid values were higher in those with more met-s traits, a phenomenon also noted in other studies. CaOx stones were less frequent in those with the most met-s traits and there was a non-significant trend to fewer CaP stones in those with more met-s traits which could also be a consequence of a lower urine pH. Urine SSUA and SSCaP mirrored these observations remaining as significant trends while SSCaOx showed no relationship to met-s traits.

## LIMITATIONS

Using registry information has many limitations that can affect both accuracy and interpretation. In this study these include diagnoses listed on problem lists which are provider dependent, variation in definition and lack of measurement of some criteria for the metabolic syndrome traits such as waist/hip ratio or fasting blood sugar levels. In addition we lacked direct measurement of bicarbonate and organic acid excretion so were unable to calculate titratable acid completely. Not all had blood chemistries within 6 months of urine collection and the number for each value varied. However, we could not detect that these issues were more problematic for any of the groups in particular. Conclusions about dietary influence on these findings are open to interpretation as even in steady state, excretion reflects not just intake but absorption and in some cases metabolism. This was an observational study that allows more for hypothesis generation than testing and none of the findings can prove causality.

## CONCLUSIONS

Our study found that stone forming patients with met-s have a defined pattern of metabolic and dietary risk factors that can contribute to increased risk of stone formation including high animal protein and associated high Na intake and lower urine pH. Greater acid excretion is largely the result of dietary factors including the higher protein intake although this does not fully explain the urine pH trends. The lower urine pH and associated higher SSUA and lower SSCaP contributed to the distribution of stone composition in those with more met-s traits. Studies are needed to determine if changing specific dietary factors can reduce risk for those with met-s and kidney stones.

## Data Availability

Data from manuscript is kept on a secure password protected folder in the secure hospital computer system

## Acknowledgements

We thank Melissa Holman and the Jeffords Institute for Quality at UVMMC for support of the Stone Registry and help with data management. Also thanks to FJ Gennari MD for manuscript review.

## REFERENCES

1. Eckel RH, Grundy SM, Zimmet PZ: The Metabolic Syndrome: Lancet 365: 1415–1428, 2005

2. Wong Y, Cook P, Roderick P, Somani BK: Metabolic Syndrome and Kidney Stone Disease: A Systematic Review of the Literature: J Endourology 30(3):246–253, 2016

3. Tasian, GE, Ross ME, Song L, Sas DJ, Keren R, Denburg MR, Chu DI, Copelovitch L, Saigal CS, Furth SL: Annual Incidence of Nephrolithiasis among Children and Adults in South Carolina from 1997 to 2012: CJASN 11(3): 488–496, 2016

4. Taylor, Eric N., Stampfer Meir J., and Curhan Gary C. Diabetes mellitus and the risk of nephrolithiasis. Kidney Int 68.3:1230–1235, 2005

5. Ferraro, Pietro M., et al. History of kidney stones and the risk of coronary heart disease: JAMA 310.4: 408–415, 2013

6. Liu Y-T, Yang P-Y, Yang Y-W, Sun H-Y, Lin I-C: The association of nephrolithiasis with metabolic syndrome and its components: a cross-sectional analysis. Ther Clin Risk Manag. 13: 41–48, 2017

7. Lutsey PL, Steffen LM, Stevens J: Dietary Intake and the Development of the Metabolic Syndrome. The Atherosclerosis Risk in Communities Study. Circulation 117:754–761, 2008

8. Adeva MM, Souto G. Diet-induced metabolic acidosis: Clin Nutr 30:416–421, 2011

9. Maalouf NM., Cameron MA, Moe OW, Adams-Huet B, Sakhaee K. Low Urine pH: A Novel Feature of the Metabolic Syndrome. CJASN 2: 883–888, 2007

10. Abate N, Chandalia M, Cabo-Chan AV, Moe OW, Sakhaee K. The metabolic syndrome and uric acid nephrolithiasis: novel features of renal manifestation in insulin resistance. Kidney Int. 65: 386–392, 2004

11. Bobulescu A, Park SK, Xu R et al. Net Acid Excretion and Urinary Organic Anions in Idiopathic Uric Acid Nephrolithiasis. CJASN 14: 1–10, 2019

12. Kok DJ, Poindexter J, Pak Charles YC: Calculation of titratable acidity from urinary stone risk factors. Kidney Int 44.1: 120–126. 1993

13. Maroni BJ, Steinman TI. Mitch WE. A Method for Estimating Nitrogen Intake of Patients with Chronic Renal Failure. Kidney Int 27.1: 58–65, 1985

14. Oh, MS: A new method for estimating G-I absorption of alkali. Kidney Int. 36: 915–917, 1989

15. Cupisti A, Meola M, D’Alessandro C, Bernabini G, Pasquali E, Carpi A, Barsotti G: Insulin resistance and low urinary citrate excretion in calcium stone formers. Biomed Pharmacother 61(1):86–90, 2007

16. Kamel KS, Cheema-Dhadli S, Halperin ML: Studies on the pathophysiology of the low urine pH in patients with uric acid stones. Kidney Int. 61: 988–994, 2002

17. Maalouf NM: Metabolic syndrome and the Genesis of Uric Acid Stones. J Ren Nutr 21(1):128–131, 2011

