## SupplementaryTable for "Association of Urine Findings with Metabolic Syndrome Traits in a Population of Patients with Nephrolithiasis"

SUPPLEMENTAL MATERIAL

**Table 1S: Supplement to Table 3**
**Comparison of Urine Variables and Metabolic Syndrome Traits** [1,2]

| | Adjusted Means | | | | |
|---|---|---|---|---|---|
| | Number of Metabolic Syndrome Traits | | | | Trend |
| | 0 | 1 | 2 | 3,4 | P Value |
| **Adjusted for age** | | | | | |
| Vol 24 (L/d) | 2.0 | 1.9 | 2.0 | 2.0 | 0.77 |
| Cr24 (mg/d) | 1330 | 1469 | 1453 | 1518 | <0.001 |
| pH | 6.1 | 6.0 | 6.0 | 5.8 | <0.001 |
| Ca24 (mg/d) | 204 | 220 | 213 | 215 | 0.44 |
| SSCaP | 1.2 | 1.2 | 1.1 | 1.0 | 0.01 |
| SSCaOx | 6.4 | 6.9 | 6.6 | 6.5 | 0.98 |
| Ox24 (mg/d) | 35 | 39 | 41 | 42 | <0.001 |
| Cit24 (mg/d) | 578 | 606 | 626 | 664 | 0.01 |
| UA24 (g/d) | 0.59 | 0.64 | 0.67 | 0.70 | <0.001 |
| SSUA | 0.8 | 1.0 | 1.1 | 1.3 | <0.001 |
| Na24 (mmol/d) | 151 | 180 | 177 | 183 | <0.001 |
| K24 (mmol/d) | 61 | 64 | 66 | 67 | 0.003 |
| Cl24 (mmol/d) | 151 | 178 | 178 | 187 | <0.001 |
| P24 (g/d) | 0.89 | 0.97 | 0.97 | 1.00 | 0.001 |
| Sul24 (mEq/d) | 37 | 40 | 39 | 41 | 0.02 |
| Mg24 (mg/d) | 99 | 103 | 104 | 106 | 0.06 |
| UUN24 (g/d) | 10.4 | 11.3 | 11.3 | 12.0 | <0.001 |
| Protein (calculated) | 80 | 88 | 88 | 95 | <0.001 |
| NH424 (mmol/d) | 33 | 36 | 36 | 38 | <0.001 |
| TAP[3] | 17 | 19 | 20 | 22 | <0.001 |
| eNAE[3] | 44 | 50 | 52 | 56 | <0.001 |
| NH424/eNAE | 0.77 | 0.70 | 0.67 | 0.61 | 0.08 |
| GI alkali[3] | 27 | 29 | 27 | 25 | 0.23 |

| | | | | | |
|---|---|---|---|---|---|
| **Adjusted for age and protein** | | | | | |
| pH | 6.1 | 6.0 | 6.0 | 5.8 | <0.001 |
| Ca24 (mg/d) | 213 | 214 | 206 | 198 | 0.09 |
| Ox24 (mg/d) | 36 | 38 | 40 | 40 | 0.02 |
| Cit24 (mg/d) | 595 | 595 | 615 | 632 | 0.21 |
| UA24 (g/d) | 0.62 | 0.63 | 0.65 | 0.65 | 0.02 |
| Na24 (mmol/d) | 160 | 175 | 171 | 167 | 0.29 |
| K24 (mmol/d) | 64 | 62 | 64 | 62 | 0.68 |
| Cl24 (mmol/d) | 159 | 174 | 173 | 172 | 0.03 |
| P24 (g/d) | 0.95 | 0.94 | 0.94 | 0.91 | 0.06 |
| Sul24 (mEq/d) | 39 | 38 | 37 | 36 | <0.001 |
| UUN24 (g/d) | 11.1 | 10.8 | 10.8 | 10.6 | <0.001 |
| NH424 (mmol/d) | 35 | 35 | 35 | 34 | 0.93 |
| TAP[3] | 18 | 19 | 19 | 19 | 0.04 |
| eNAE[3] | 47 | 48 | 50 | 50 | 0.06 |
| NH424/eNAE | 0.76 | 0.71 | 0.68 | 0.63 | 0.17 |
| GI alkali[3] | 28 | 28 | 26 | 23 | 0.01 |
| **Adjusted for age and sulfate** | | | | | |
| pH | 6.1 | 6.0 | 6.0 | 5.8 | <0.001 |
| Ca24 (mg/d) | 209 | 216 | 213 | 209 | 0.91 |
| Ox24 (mg/d) | 36 | 38 | 41 | 42 | <0.001 |
| Cit24 (mg/d) | 586 | 596 | 626 | 651 | 0.03 |
| UA24 (g/d) | 0.60 | 0.63 | 0.67 | 0.68 | <0.001 |
| Na24 (mmol/d) | 155 | 176 | 176 | 177 | <0.001 |
| K24 (mmol/d) | 63 | 62 | 66 | 65 | 0.04 |
| Cl24 (mmol/d) | 155 | 175 | 178 | 182 | <0.001 |
| P24 (g/d) | 0.92 | 0.94 | 0.97 | 0.96 | 0.02 |
| Sul24 (mEq/d) | -- | -- | -- | -- | -- |
| UUN24 (g/d) | 10.8 | 10.9 | 11.2 | 11.5 | <0.001 |
| NH424 (mmol/d) | 34 | 35 | 36 | 37 | 0.01 |
| TAP[3] | 18 | 19 | 20 | 21 | <0.001 |
| eNAE[3] | 46 | 48 | 52 | 54 | <0.001 |
| NH424/eNAE | 0.77 | 0.71 | 0.67 | 0.62 | 0.11 |
| GI alkali[3] | 28 | 28 | 27 | 24 | 0.07 |

| | | | | | |
|---|---|---|---|---|---|
| **Adjusted for age and GI alkali** | | | | | |
| pH | 6.1 | 6.0 | 6.0 | 5.8 | <0.001 |
| Ca24 (mg/d) | 204 | 218 | 214 | 219 | 0.24 |
| Ox24 (mg/d) | 35 | 39 | 41 | 42 | <0.001 |
| Cit24 (mg/d) | 578 | 597 | 630 | 678 | <0.001 |
| UA24 (g/d) | 0.59 | 0.64 | 0.67 | 0.70 | <0.001 |
| Na24 (mmol/d) | 151 | 178 | 177 | 186 | <0.001 |
| K24 (mmol/d) | 61 | 63 | 66 | 69 | <0.001 |
| Cl24 (mmol/d) | 151 | 177 | 179 | 189 | <0.001 |
| P24 (g/d) | 0.90 | 0.97 | 0.97 | 1.01 | <0.001 |
| Sul24 (mEq/d) | 37 | 40 | 39 | 41 | 0.008 |
| UUN24 (g/d) | 10.4 | 11.2 | 11.3 | 12.1 | <0.001 |
| NH424 (mmol/d) | 33 | 36 | 36 | 37 | 0.001 |
| TAP[3] | 17 | 19 | 20 | 21 | <0.001 |
| eNAE[3] | 44 | 50 | 52 | 55 | <0.001 |
| NH424/eNAE | 0.77 | 0.71 | 0.67 | 0.61 | 0.07 |
| GI alkali[3] | -- | -- | -- | -- | -- |

Table 2S: Metabolic Syndrome Correlation Coefficients

| | Protein | Age | Cit24 | Na24 | K24 | Nh424 | GIalkali |
|---|---|---|---|---|---|---|---|
| Protein | 1 | | | | | | |
| Age | 0.03 | 1 | | | | | |
| P value | 0.19 | | | | | | |
| Cit24 | 0.26 | 0.06 | 1 | | | | |
| P value | <0.001 | 0.02 | | | | | |
| Na24 | 0.61 | -0.07 | 0.24 | 1 | | | |
| P value | <0.001 | 0.01 | <0.001 | | | | |
| K24 | 0.60 | 0.16 | 0.38 | 0.43 | 1 | | |
| P value | <0.001 | <0.001 | <0.001 | <0.001 | | | |
| Nh424 | 0.60 | -0.17 | 0.08 | 0.32 | 0.15 | 1 | |
| P value | <0.001 | <0.001 | 0.004 | <0.001 | <0.001 | | |
| GIalkali | 0.22 | 0.09 | 0.39 | 0.39 | 0.54 | -0.32 | 1 |
| P value | <0.001 | <0.001 | <0.001 | <0.001 | <0.001 | <0.001 | |
